# Serum VEGF Levels on Admission in COVID-19 Patients Correlate with SP-D and Neutrophils, Reflecting Disease Severity: A Prospective Study

**DOI:** 10.1101/2023.07.17.23292653

**Authors:** Mayoko Tsuji, Mitsuko Kondo, Yasuto Sato, Azusa Miyoshi, Fumi Kobayashi, Ken Arimura, Kaoru Yamashita, Satoshi Morimoto, Naoko Yanagisawa, Atsuhiro Ichihara, Etsuko Tagaya

**Author notes:** **Corresponding author details:** Name: Mayoko Tsuji, Twitter account name (optional): @May_resp.

## Abstract

**Background and objective:** The COVID-19 pandemic caused by SARS-CoV-2 has resulted in significant global morbidity and mortality. This study aimed to investigate the clinical significance of serum vascular endothelial growth factor (VEGF) in COVID-19 patients and its association with disease severity and pulmonary injury.

**Methods:** We prospectively collected data from 71 hospitalized COVID-19 patients between June 2020 and January 2021. Patients were classified as either mild or severe based on their oxygen requirements during hospitalization. Serum VEGF levels were measured using an ELISA kit.

**Results:** In comparison to mild cases, significantly elevated serum VEGF levels were observed in severe COVID-19 patients. Furthermore, VEGF levels exhibited a positive correlation with white blood cell count, neutrophil count, and lymphocyte count. Notably, serum surfactant protein-D (SP-D), an indicator of alveolar epithelial cell damage, was significantly higher in patients with elevated VEGF levels.

**Conclusion:** These results suggest that elevated serum VEGF levels could serve as a prognostic biomarker for COVID-19 as it is indicative of alveolar epithelial cell injury caused by SARS-CoV-2 infection. Additionally, we observed a correlation between VEGF and neutrophil activation, which plays a role in the immune response during endothelial cell injury, indicating a potential involvement of angiogenesis in disease progression. Further research is needed to elucidate the underlying mechanisms of VEGF elevation in COVID-19.

## INTRODUCTION

COVID-19, caused by SARS-CoV2, was first reported in 2019 in China ^1^, and rapidly escalated into a global pandemic. This unprecedented crisis has resulted in approximately 7 million deaths worldwide, according to the World Health Organization (https://covid19.who.int/). The primary target of SARS-CoV2 infection is the alveolar type LJ epithelial cells (ATLJ). Infection of these cells can lead to cell death through apoptosis or pyroptosis mechanisms ^2–4^. However, severe endothelial injury with intracellular viral presence has been observed in the lungs of patients with COVID-19^5–7^.

Angiogenesis, the process involving new blood vessel formation from pre-existing vessels, is a fundamental process involved in various conditions such as cancer, wound healing, and multiple myeloma ^8^. There are two described types of angiogenesis, intussusceptive and sprouting angiogenesis ^9–12^. Notably, intussusceptive angiogenesis has been identified as occurring in COVID-19 autopsy lung samples, and its occurrence appears to be more prominent in COVID-19 compared to influenza virus pneumoniae ^6^.

COVID-19 severity has been associated with vascular complications such as cardiac failure, renal failure, coagulopathy, stroke, and pulmonary embolism ^13, 14^. Also, severe pneumonia often progresses to a life threatening condition known as acute respiratory distress syndrome (ARDS) ^15, 16^. The development of ARDS is attributed to endothelial cell and epithelial cell damage^17^. Collectively, these observations highlight the critical role of vasculopathy in the pathogenesis of COVID-19.

The serum level of vascular endothelial growth factor (VEGF) has been found to exhibit a positive correlation with disease severity in COVID-19 patients. VEGF is recognized as an angiocrine factor and holds potential as a prognostic marker for COVID-19 ^18, 19^. However, the precise implications of VEGF in the pathophysiology of COVID-19 remain unclear, and its clinical significance in the context of the disease is yet be determined.

In this study, we aimed to explore and investigate the clinical relevance of serum VEGF levels in COVID-19 patients.

## METHODS

### Study design and population

The protocol for this study was approved by the ethical committee of Tokyo Women’s Medical University (TWMU), and registered at Clinical Trials (No. 5612-R). All participants gave written informed consent.

We prospectively collected data from patients admitted and hospitalized with COVID-19 at TWMU hospital between June, 2020 and January, 2021. The cohort included all symptomatic patients who tested positive with SARS-CoV2 through PCR testing, resulting in a total of 71 confirmed cases. The severity of the disease was categorized according to the oxygen requirements upon admission, with patients either classified as mild (O_2_ intake < 3 L/min; n=46, 64.8%) or severe (O_2_ intake ≧3 L/min; n =25, 35.2%).

Patients in this study were individuals who visited the Fever Outpatient Department in TWMU, and were subsequently recommended for hospitalization by Tokyo Center for Infectious Disease Control and Prevention (iCDC).

Demographic data, including age, sex, BMI, disease comorbidity, oxygen level, history of smoking, history of alcohol, blood cell counts, serum KL-6, serum SP-D, serum D-dimer, and serum sIL-2R levels were obtained from medical records and medical history forms. Serum VEGF levels were measured using ELISA kit.

### Analysis of serum VEGF

To measure the serum VEGF levels, plasma samples were collected at the time of hospitalization. We utilized an enzyme-linked immunosorbent assay (ELISA) kit (Ray Biotech ELH-VEGF) in duplicate, and the absorbance was measured at a wave length of 450 nm using an Microplate Reader (Bio-Rad 680).

### Outcomes

The primary objectives of this study were to explore the pathogenetic and clinical significance of serum VEGF by comparing clinical findings in severe COVID-19 patients.

### Statistical Analysis

Continuous variables were presented as mean ±LJstandard error of the mean (SEM), while categorical variables were expressed as numbers (proportion) in patients’ characteristics. Continuous variables between the 2 groups were compared using the Mann-Whitney U test and Fisher’s exact test was used for categorical variables. Bivariate correlations were calculated using Spearman’s rank correlation coefficient between continuous biological variables and ordinal variables. *p* <0.05 was considered as statistically significant. *r* =0.2 -0.39 was considered as mild correlation, and *r* =0.4 -0.69 was considered as moderate correlation.

For statistical analysis, the data were analyzed using the Prism 9 software package (GraphPad Software, San Diego, CA, USA).

## RESULTS

### Patients’ characteristics

During the study period, a total of 71 patients (50 men and 21 women) with a mean of age of 58.5 years (range 49-69) who were hospitalized due to COVID-19 agreed to participate in this research. All 71 patients had undergone SARS-CoV-2 PCR testing at least one day before admission, confirming a positive result. The severity of COVID-19 was determined based on the maximum inhaled oxygen flow during hospitalization, categorizing patients as either Mild, < 3 L/min (n = 46) or Severe, ≧ 3 L/min (n = 25). The majority of patients in the study were male (Mild: 60.9%, Severe: 88%, *p* =0.028), with smoking history more prevalent among the mild group (Mild: 71.1%, Severe: 56.0%, *p* = 0.29), while habitus of alcohol did not significantly differ between the groups (Mild: 63.0%, Severe: 64.0%, *p* >0.999). Differences between Mild group and Severe group were observed in terms of age (range; Mild: 49-69, Severe: 49-65, *p* = 0.052) and BMI (range; Mild: 20.3-27.4, Severe: 22.8-30.1, *p* = 0.0087). Regarding comorbidities, the frequencies of hypertension (Mild: 25.0%, Severe: 68.0%, *p* =0.0008), chronic kidney disease (Mild: 13.0%, Severe: 44.0%, *p* =0.0074), and kidney transplantation (Mild: 9.1%, Severe: 66.7%, *p* =0.023) were significantly different between the mild and severe group (Table 1).

**Table 1.** Clinical characteristics of COVID-19 patients. Data are presented as Median (IQR) or n (%). Statistical analysis was performed by ordinary Mann Whitney’s U test. \**p* < 0.05.

| Variable | Mild(n=46) | Severe(n=25) | P value |
| --- | --- | --- | --- |
| **Age** |  |  |  |
| Median (IQR) | 58.5(49-69) | 66(49-65) | 0.052 |
| **Sex** |  |  |  |
| Male n (%) | 28(60.9%) | 22(88.0%) | 0.028* |
| **BMI** |  |  |  |
| Median (IQR) | 23.0(20.3-27.4) | 26.1(22.8-30.1) | 0.0087* |
| **Smoking history** | 32(71.1%) | 14(56.0%) | 0.29 |
| **Alcohol** | 29(63.0%) | 16(64.0%) | >0.99 |
| **Comorbidities** |  |  |  |
| Cancer | 6(13.0%) | 4(16%) | 0.73 |
| Diabetes | 9(19.6%) | 9(36.0%) | 0.16 |
| Hypertension | 11(25.0%) | 17(68.0%) | 0.0008* |
| Dyslipidemia | 13(29.6%) | 13(52.0%) | 0.076 |
| Heart failure | 0(0.0%) | 2(8.0%) | 0.13 |
| Chronic kidney disease | 6(13.0%) | 11(44.0%) | 0.0074* |
| Kidney transplantation | 4(9.1%) | 8(66.7%) | 0.023* |
| Chronic liver disease | 6(13.0%) | 2(8.0%) | 0.7 |
| Hemodialysis | 1(2.2%) | 2(2.8%) | 0.282 |
| **FiO2** |  |  |  |
| Median(IQR) | 21(21-24) | 40(32-100) | <0.0001* |

### Prognosis markers of COVID-19 on admission

To identify prognostic markers of COVID-19, we analyzed multiple factors in serum samples obtained from COVID-19 patients on admission. Initially, we examined endothelial injury markers, such as VEGF and D-dimer. VEGF levels were significantly elevated in the Severe group (*p* =0.027) (Figure 1a). Moreover, WBC counts (*p* =0.00040) and neutrophil counts (*p* <0.0001) (Figure 1e and f) were significantly higher in severe COVID-19 patients. As sIL-2R stimulates lymphocytes, Conversely, there were no significant differences in KL-6 (*p* = 0.30) and SP-D levels (*p* = 0.50), which are indicators of alveolar epithelial damage (Figure 1c and d).

**Figure 1.**
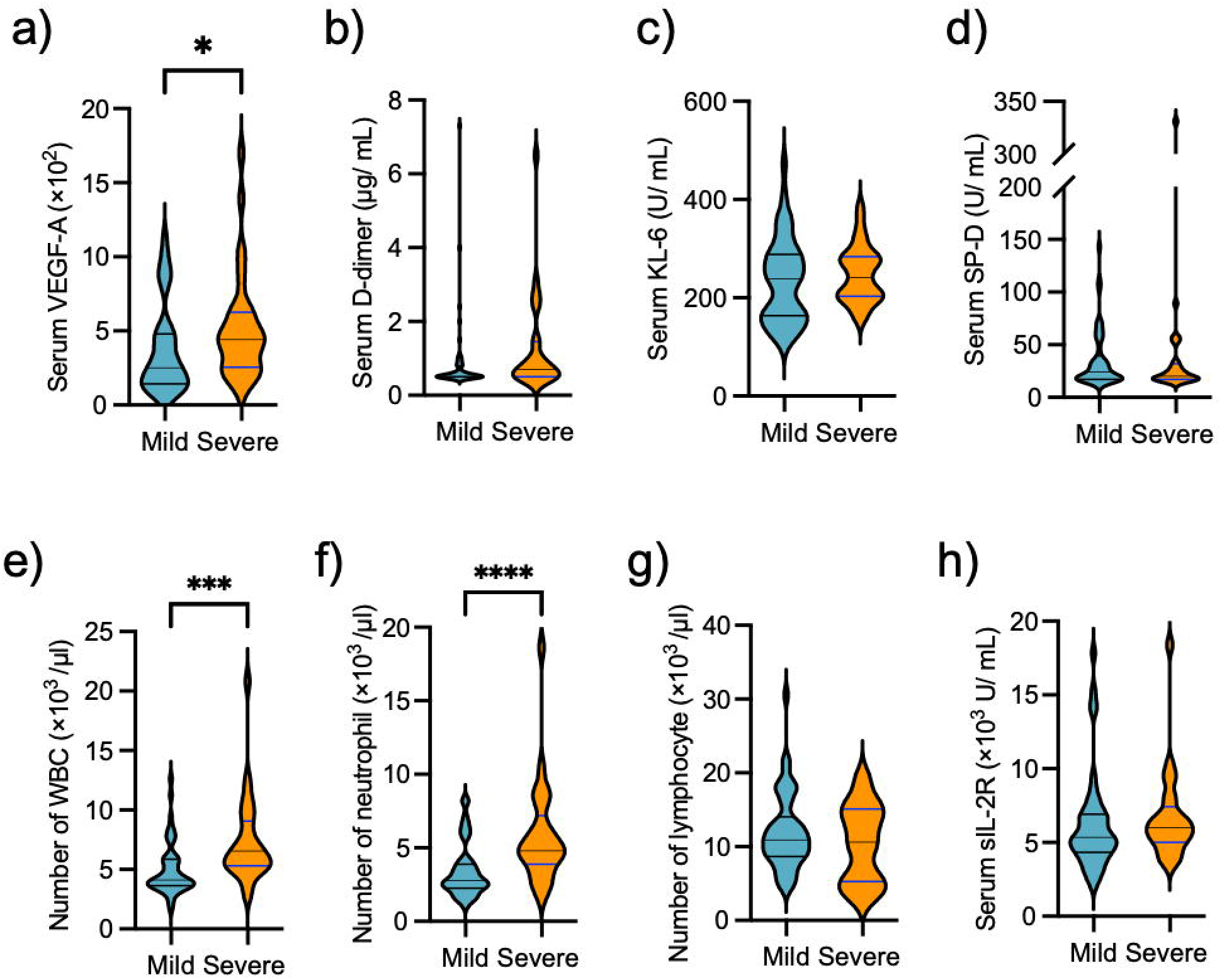
Serum endothelial cell and alveolar epithelial cell damage markers of COVID-19 patients on admission. Mild (n=46) and Severe(n=25). Statistical analysis was performed by ordinary Mann–Whitney’s U test. VEGF-A: vascular endothelial growth factor, WBC: white blood cell. \**p* < 0.05, ** *p* < 0.01, *** *p* < 0.001, **** *p* < 0.0001.

We then focused on exploring the relationship between serum VEGF levels and disease severity. ROC analysis demonstrated an area under the curve (AUC) of 0.68 [95%CI: 0.57-0.82], with a cut off value of 394 pg/μl (sensitivity: 72%, Specificity: 65%) determined using the Youden Index (Figure 2).

**Figure 2.**
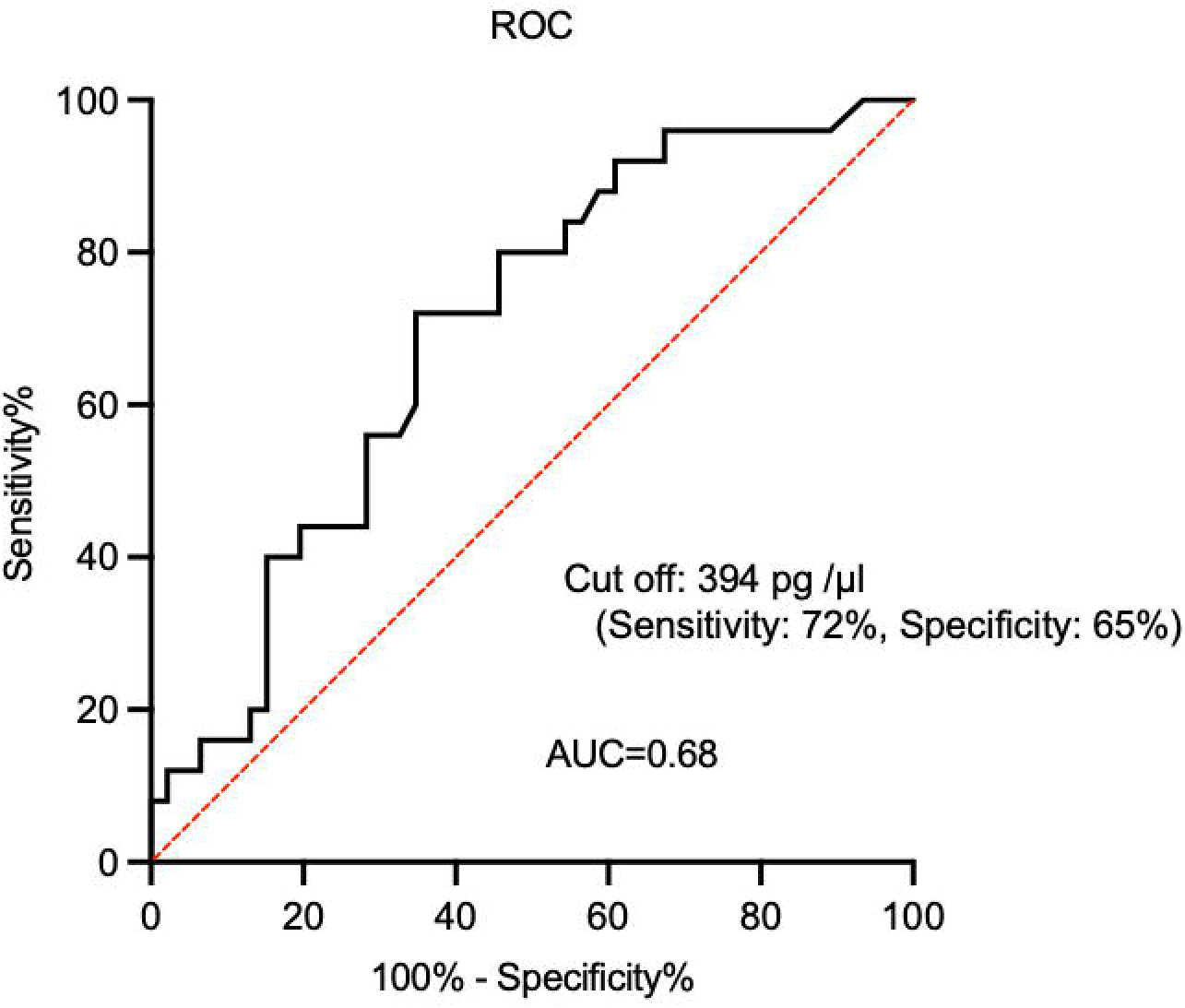
ROC analysis of Severity and VEGF.

### Correlation between VEGF and SP-D in VEGF high patients

As VEGF is commonly produced by epithelial cells, we next examined the relationship between VEGF and epithelial damage markers such as SP-D. Patients with serum VEGF levels >394 pg/μl were classified as the VEGF high group, based on the identified cut off value for prognosis of COVID-19 severity. One patient with interstitial pneumoniae was excluded from the analysis (n= 33). Bivariate correlations using Spearman’s rank correlation coefficient revealed a positive correlation between VEGF and SP-D in the VEGF high group (*r* = 0.42, *p* = 0.015) (Figure 3a), while no significant correlation was observed between VEGF and KL-6 levels (*r* = -0.058, *p* = 0.75) (Figure 3b). This data suggests that the VEGF high group of COVID-19 patients exhibits alveolar damage.

**Figure 3.**
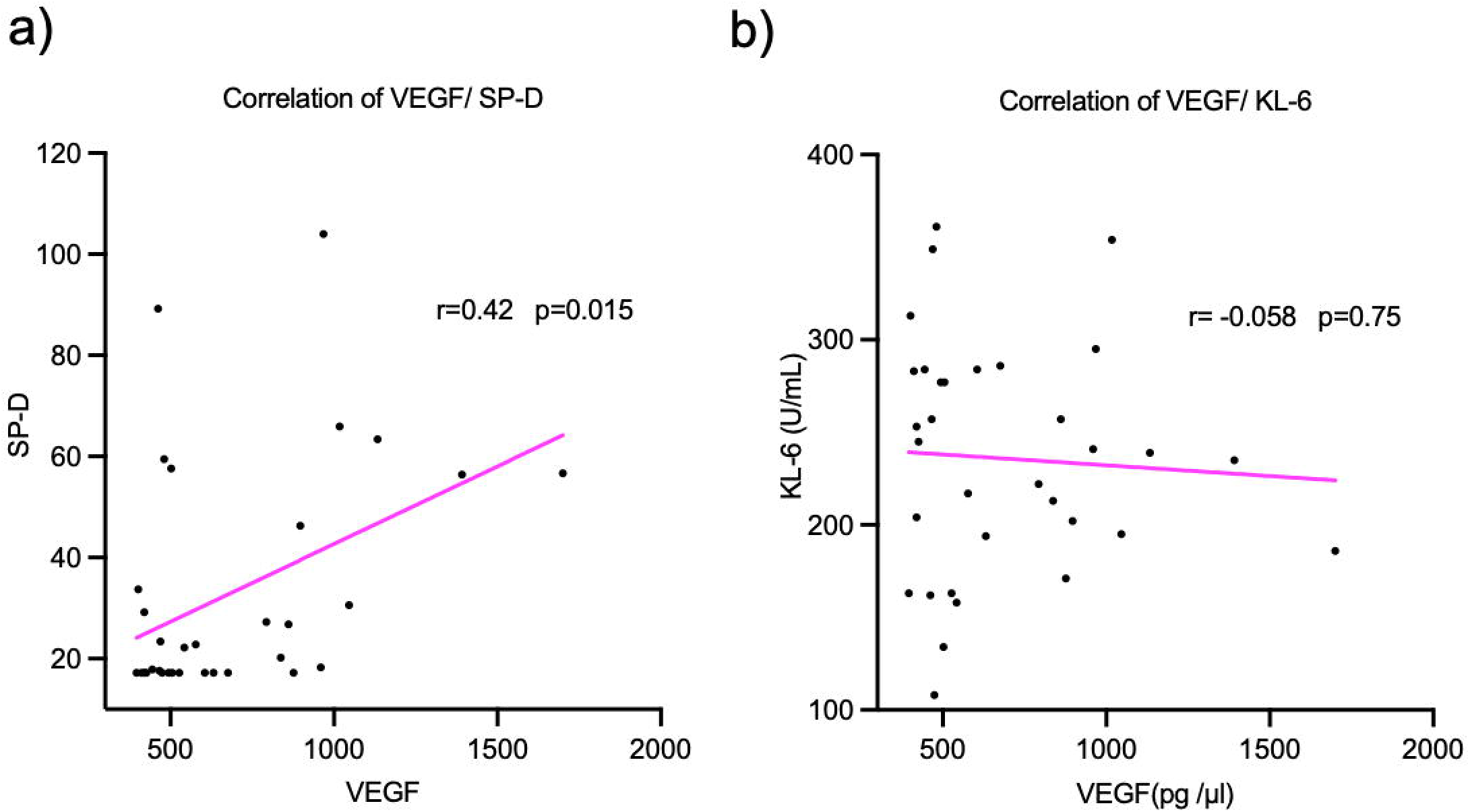
VEGF correlates to acute alveolar damage in severe prognosis patients.

### Correlation between VEGF and Neutrophils

Our data showed that WBC and Neutrophil levels were higher in the Severe group (Figure 1). We assessed correlations between VEGF and various markers using bivariate analysis. We observed correlations between VEGF and WBC (r= 0.36, *p*= 0.0019) (Figure 4a), Neutrophil (r= 0.34, *p*= 0.0041) (Figure 4b), and Lymphocyte (r= 0.26, *p*= 0.030) (Figure 4c).

**Figure 4.**
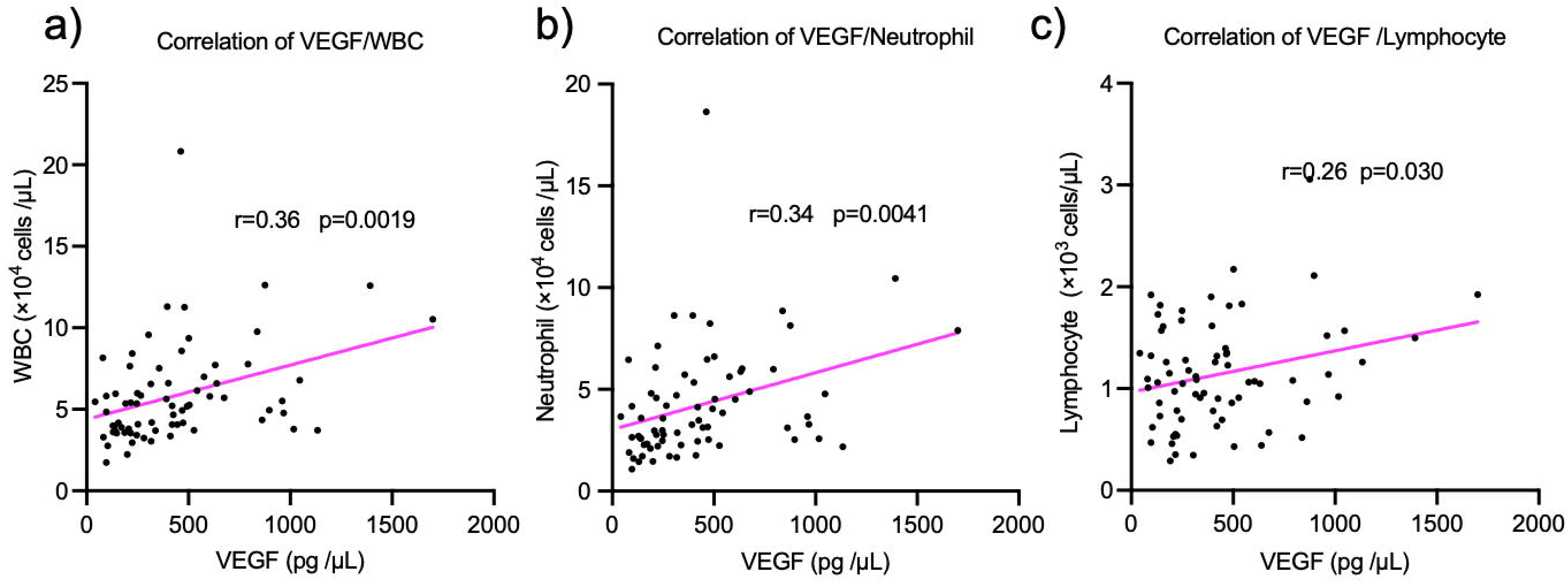
Correlations between VEGF and biological variables (white blood cell count, neutrophil, and lymphocyte) using Pearson’s correlation coefficient. WBC: white blood cell, VEGF: vascular endothelial growth factor.

## DISCUSSION

Our prospective study revealed that serum VEGF levels served as a prognostic marker for COVID-19. These levels were correlated with white blood cell counts, neutrophil counts, and lymphocyte counts at the time of hospitalization. Notably, among patients with elevated VEGF levels (VEGF > 394 pg/μl), we observed a significant correlation with serum SP-D. This indicates a potential correlation between VEGF elevation and alveolar epithelial cell injury caused by SARS-CoV2 infection.

In this study, patient characteristics revealed a higher proportion of males, smokers, and individuals with a history of alcohol consumption (Table1) compared to general public population. This observation can be attributed to the proximity of our study site to downtown areas. Furthermore, our data demonstrated elevated levels of WBCs, neutrophils, and VEGF at the time of hospitalization, which were associated with the severity of COVID-19 (Figures 1 and 4). Additionally, the severe COVID-19 group exhibited a significantly higher prevalence of comorbidities such as hypertension, chronic kidney disease, kidney transplantation (Table 1), and uric acid (dana not shown ^20^). Based on our patient characteristics and biological markers, these findings suggest that factors contributing to disease severity are consistent with previous public reports and risk assessment tests ^21, 22^. These results reinforce the consistency between our patient characteristics and biological markers in relation to disease severity. Furthermore, we conducted further investigation of these factors among both male and female patients. Interestingly, we observed the same trends in both genders, with a positive correlation between VEGF levels and disease severity. The trends displayed moderate correlation in females (*r* =0.57, *p* =0.0067), and mild correlation in males (*r* =0.39, *p* =0.0052) (Supplemental figure 1). Collectively, our comprehensive analysis, considering patient characteristics and relevant biological markers, suggests that the severity factors observed in our study align with those reported in the general population and are consistent with established risk assessment tests.

Epithelial cell injury is a common feature observed in various respiratory diseases including infectious pneumonia, ARDS, and interstitial pneumonia. Such injury results in hypoxia due to inadequate oxygen uptake. In COVID-19 patients, SARS-CoV2 infects alveolar type II cells (ATII) expressing ACE2, leading to apoptosis and pyroptosis of the infected cells ^23, 24^. Subsequently, alveolar macrophages are activated and release proinflammatory cytokines following SARS-Cov2 infection ^25, 26^. These cytokines contribute to endothelial cell dysfunction, ultimately leading to angiogenesis ^27, 28^. In this study, we have identified a correlation between VEGF and SP-D (Figure 3), suggesting that elevated VEGF levels reflects epithelial cell injury, particularly in patients with high VEGF levels. Furthermore, activated endothelial cells stimulate neutrophils and promote platelet accumulation by releasing RANTES and PF4 ^29, 30^. Our data demonstrated that VEGF, an indicator of angiogenesis, was correlated with WBC and neutrophil counts (Figure 1e and f). Notably, among patients with high WBC and neutrophils, only one patient developed bacterial pneumonia as a complication. This finding suggests a stronger association between the effect of COVID-19 and less association with complications such as bacterial infection, which typically exhibit elevated WBC and neutrophil counts.

Serum SP-D is known to rapidly increase when ATII cell damage occurs, as seen in interstitial lung disease, and its level at the initial visit is considered a poor prognostic marker ^31^. KL-6, on the other hand, also reflects ATII cell damage but is not specific to alveolar epithelial cells and increases later in response to cell damage ^32, 33^ due to its higher molecular weight. In our data, SP-D levels but not KL-6 levels at the time of hospitalization was elevated in patients with high levels of VEGF (Figure 3a and 3b). This discrepancy may be attributed to the fact that our samples were obtained during the early stage of infection. The correlation between VEGF and SP-D suggests an interaction between epithelial cells and endothelial cells.

Peng Wang et al., reported that “vasculature development related proteins” were upregulated in SARS-CoV2-infected alveolar epithelial cells *in vitro*. Additionally, “neutrophil activation related proteins” were enriched in SARS-CoV2-infected endothelial cells, leading to neutrophil activation, while mediated immunity-related proteins were also enriched^34^. Our data further support the cascades of SARS-CoV2 mediated neutrophil activation via angiogenesis initiated by alveolar epithelial cells in COVID-19 patients.

VEGF is known to be produced by perivascular cells, including epithelial cells, neutrophils, and mesenchymal cells, in response to injury triggered by hypoxia. It is involved in production of MMP-9 and plays a role in angiogenesis in tumors ^35–37^. Our data revealed an increase in oxygen demand that correlated with serum VEGF levels (Table 1), suggesting that hypoxia may be one of the factors contributing to VEGF upregulation.

It is worth noting that our samples were obtained from patients infected with the Delta variant, which is known to be associated with a high risk of endothelial dysfunction. Although the severity of cases after the Omicron variant is reported to be less critical, some severe patients still exhibit vascular defects. Further data analysis following Omicron variant infection is warranted.

In conclusion, our findings demonstrate a correlation between serum VEGF levels and neutrophil counts in COVID-19 patients. Specifically, patients with remarkably high levels of VEGF showed a correlation with SP-D. These observations suggest that the pathogenesis of SARS-CoV2-induced alveolar endothelial cell and endothelial cell damage subsequently activates neutrophils. VEGF, as a prognostic marker for COVID-19, serves as an indicator of alveolar epithelial cell and endothelial cell damage, as well as neutrophilic inflammation. These findings contribute to our understanding of the complex interplay between VEGF, cellular damage, and immune response in the context of COVID-19.

## Data Availability

All data produced in the present study are available upon reasonable request to the authors

## Acknowledgements

The authors thank Atsuko Ohnishi, Masayuki Shino, and Yoshimi Sugimura for their technical support. This work was supported by a research grant from the TWMU Career Development Center for Medical Professionals. And we thank Shigeru Ashino from the Department of Microbiology and Immunology, TWMU, for technical support on the environment of the biosafety-level laboratory.

## Human/Animal Ethics Approval Declaration

The protocol was approved by the ethical committee, Tokyo Women’s Medical University (TWMU), and registered at Clinical Trials (No. 5612-R). All participants gave written informed consent.

---

**Supplemental Figure 1.**
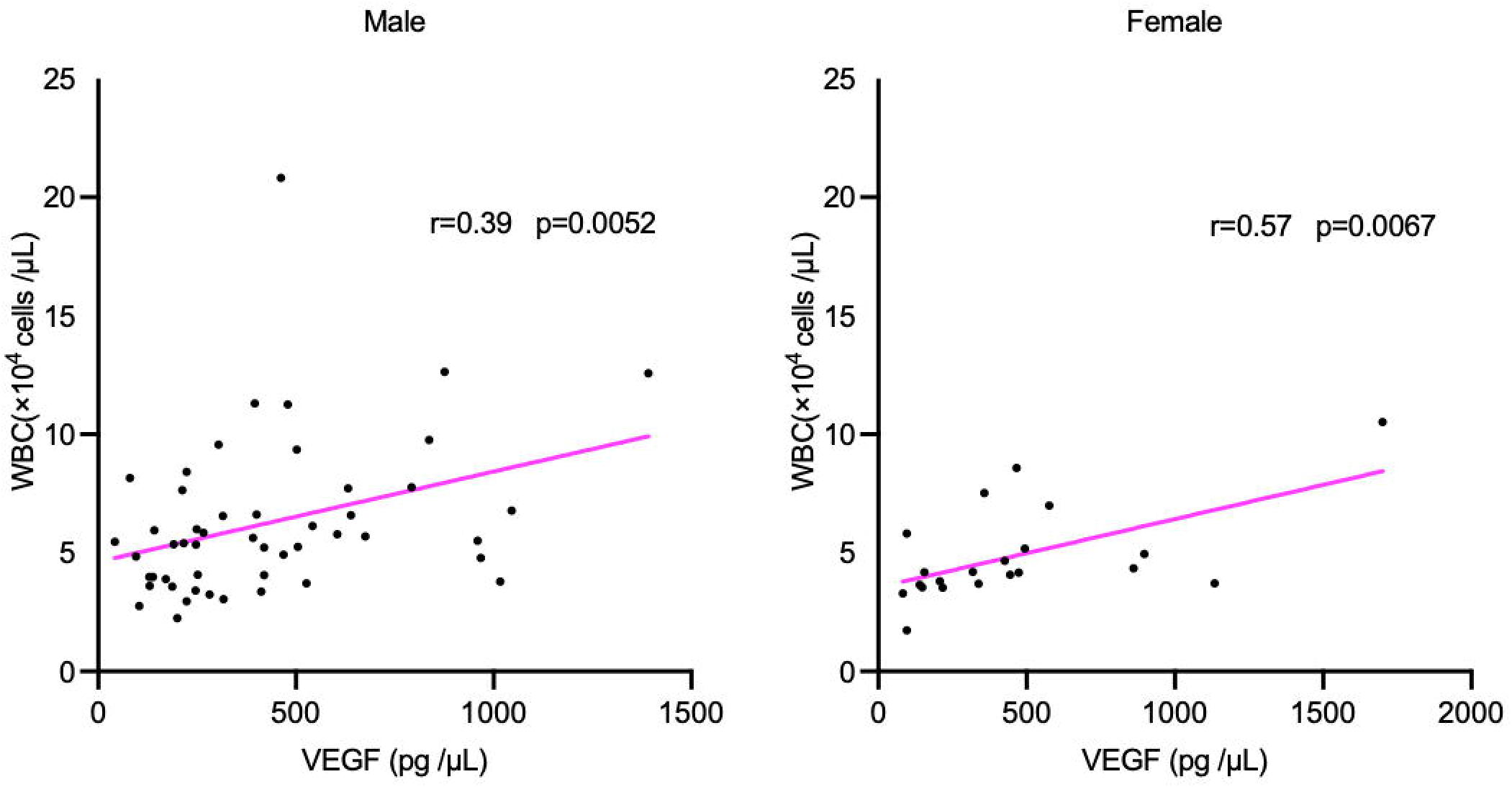
Correlations between serum VEGF and WBC by sex. Serum VEGF and WBC were correlated in both Male and Female .

## REFERENCES

1 Zhu N, Zhang D, Wang W, Li X, Yang B, Song J, Zhao X, Huang B, Shi W, Lu R, Niu P, Zhan F, Ma X, Wang D, Xu W, Wu G, Gao GF, Tan W, Team CNCIaR. A Novel Coronavirus from Patients with Pneumonia in China, 2019. N Engl J Med. 2020; 382: 727–33.

2 Mossel EC, Wang J, Jeffers S, Edeen KE, Wang S, Cosgrove GP, Funk CJ, Manzer R, Miura TA, Pearson LD, Holmes KV, Mason RJ. SARS-CoV replicates in primary human alveolar type II cell cultures but not in type I-like cells. Virology. 2008; 372: 127–35.

3 Qian Z, Travanty EA, Oko L, Edeen K, Berglund A, Wang J, Ito Y, Holmes KV, Mason RJ. Innate immune response of human alveolar type II cells infected with severe acute respiratory syndrome-coronavirus. Am J Respir Cell Mol Biol. 2013; 48: 742–8.

4 Wang P, Luo R, Zhang M, Wang Y, Song T, Tao T, Li Z, Jin L, Zheng H, Chen W, Zhao M, Zheng Y, Qin J. A cross-talk between epithelium and endothelium mediates human alveolar-capillary injury during SARS-CoV-2 infection. Cell Death Dis. 2020; 11: 1042.

5 Ahmetaj-Shala B, Peacock TP, Baillon L, Swann OC, Gashaw H, Barclay WS, Mitchell JA. Resistance of endothelial cells to SARS-CoV-2 infection *in vitro*. bioRxiv. 2020: 2020.11.08.372581.

6 Ackermann M, Verleden SE, Kuehnel M, Haverich A, Welte T, Laenger F, Vanstapel A, Werlein C, Stark H, Tzankov A, Li WW, Li VW, Mentzer SJ, Jonigk D. Pulmonary Vascular Endothelialitis, Thrombosis, and Angiogenesis in Covid-19. N Engl J Med. 2020; 383: 120–8.

7 Iwamura C, Hirahara K, Kiuchi M, Ikehara S, Azuma K, Shimada T, Kuriyama S, Ohki S, Yamamoto E, Inaba Y, Shiko Y, Aoki A, Kokubo K, Hirasawa R, Hishiya T, Tsuji K, Nagaoka T, Ishikawa S, Kojima A, Mito H, Hase R, Kasahara Y, Kuriyama N, Tsukamoto T, Nakamura S, Urushibara T, Kaneda S, Sakao S, Tobiume M, Suzuki Y, Tsujiwaki M, Kubo T, Hasegawa T, Nakase H, Nishida O, Takahashi K, Baba K, Iizumi Y, Okazaki T, Kimura MY, Yoshino I, Igari H, Nakajima H, Suzuki T, Hanaoka H, Nakada TA, Ikehara Y, Yokote K, Nakayama T. Elevated Myl9 reflects theMyl9-containing microthrombi in SARS-CoV-2-induced lung exudative vasculitis and predicts COVID-19 severity. Proc Natl Acad Sci U S A. 2022; 119: e2203437119.

8 Folkman J. Tumor angiogenesis: therapeutic implications. N Engl J Med. 1971; 285: 1182–6.

9 Bär T, Wolff JR. The formation of capillary basement membranes during internal vascularization of the rat’s cerebral cortex. Z Zellforsch Mikrosk Anat. 1972; 133: 231–48.

10 Marin-Padilla M. Early vascularization of the embryonic cerebral cortex: Golgi and electron microscopic studies. J Comp Neurol. 1985; 241: 237–49.

11 del Toro R, Prahst C, Mathivet T, Siegfried G, Kaminker JS, Larrivee B, Breant C, Duarte A, Takakura N, Fukamizu A, Penninger J, Eichmann A. Identification and functional analysis of endothelial tip cell-enriched genes. Blood. 2010; 116: 4025–33.

12 Gerhardt H, Golding M, Fruttiger M, Ruhrberg C, Lundkvist A, Abramsson A, Jeltsch M, Mitchell C, Alitalo K, Shima D, Betsholtz C. VEGF guides angiogenic sprouting utilizing endothelial tip cell filopodia. J Cell Biol. 2003; 161: 1163–77.

13 Huang C, Wang Y, Li X, Ren L, Zhao J, Hu Y, Zhang L, Fan G, Xu J, Gu X, Cheng Z, Yu T, Xia J, Wei Y, Wu W, Xie X, Yin W, Li H, Liu M, Xiao Y, Gao H, Guo L, Xie J, Wang G, Jiang R, Gao Z, Jin Q, Wang J, Cao B. Clinical features of patients infected with 2019 novel coronavirus in Wuhan, China. Lancet. 2020; 395: 497–506.

14 Smadja DM, Mentzer SJ, Fontenay M, Laffan MA, Ackermann M, Helms J, Jonigk D, Chocron R, Pier GB, Gendron N, Pons S, Diehl JL, Margadant C, Guerin C, Huijbers EJM, Philippe A, Chapuis N, Nowak-Sliwinska P, Karagiannidis C, Sanchez O, Kümpers P, Skurnik D, Randi AM, Griffioen AW. COVID-19 is a systemic vascular hemopathy: insight for mechanistic and clinical aspects. Angiogenesis. 2021; 24: 755–88.

15 Diehl JL, Peron N, Chocron R, Debuc B, Guerot E, Hauw-Berlemont C, Hermann B, Augy JL, Younan R, Novara A, Langlais J, Khider L, Gendron N, Goudot G, Fagon JF, Mirault T, Smadja DM. Respiratory mechanics and gas exchanges in the early course of COVID-19 ARDS: a hypothesis-generating study. Ann Intensive Care. 2020; 10: 95.

16 Karagiannidis C, Mostert C, Hentschker C, Voshaar T, Malzahn J, Schillinger G, Klauber J, Janssens U, Marx G, Weber-Carstens S, Kluge S, Pfeifer M, Grabenhenrich L, Welte T, Busse R. Case characteristics, resource use, and outcomes of 10LJ021 patients with COVID-19 admitted to 920 German hospitals: an observational study. Lancet Respir Med. 2020; 8: 853–62.

17 Vassiliou AG, Kotanidou A, Dimopoulou I, Orfanos SE. Endothelial Damage in Acute Respiratory Distress Syndrome. Int J Mol Sci. 2020; 21.

18 Syed F, Li W, Relich RF, Russell PM, Zhang S, Zimmerman MK, Yu Q. Excessive Matrix Metalloproteinase-1 and Hyperactivation of Endothelial Cells Occurred in COVID-19 Patients and Were Associated With the Severity of COVID-19. J Infect Dis. 2021; 224: 60–9.

19 Rovas A, Osiaevi I, Buscher K, Sackarnd J, Tepasse PR, Fobker M, Kühn J, Braune S, Göbel U, Thölking G, Gröschel A, Pavenstädt H, Vink H, Kümpers P. Microvascular dysfunction in COVID-19: the MYSTIC study. Angiogenesis. 2021; 24: 145–57.

20 Yamashita K, Morimoto S, Kimura S, Seki Y, Bokuda K, Watanabe D, Tsuji M, Arimura K, Shimamoto K, Tagaya E, Kawan M, Ichihara A. Hyperuricemia and the Severity of Coronavirus Disease 2019 in Japan: A Retrospective Cohort Study - An Inseparable Relation with Hypertension and Chronic Kidney Disease. Endocrinology, Diabetes and Metabolism Journal. May 22, 2023; **Volume** 7

21 Clift AK, Coupland CAC, Keogh RH, Diaz-Ordaz K, Williamson E, Harrison EM, Hayward A, Hemingway H, Horby P, Mehta N, Benger J, Khunti K, Spiegelhalter D, Sheikh A, Valabhji J, Lyons RA, Robson J, Semple MG, Kee F, Johnson P, Jebb S, Williams T, Hippisley-Cox J. Living risk prediction algorithm (QCOVID) for risk of hospital admission and mortality from coronavirus 19 in adults: national derivation and validation cohort study. BMJ. 2020; 371: m3731.

22 Eythorsson E, Bjarnadottir V, Runolfsdottir HL, Helgason D, Ingvarsson RF, Bjornsson HK, Olafsdottir LB, Bjarnadottir S, Agustsson AS, Oskarsdottir K, Thorvaldsson HH, Kristjansdottir G, Bjornsson AH, Emilsdottir AR, Armannsdottir B, Gudlaugsson O, Hansdottir S, Gottfredsson M, Bjarnason A, Sigurdsson MI, Indridason OS, Palsson R. Development of a prognostic model of COVID-19 severity: a population-based cohort study in Iceland. Diagn Progn Res. 2022; 6: 17.

23 Ortiz ME, Thurman A, Pezzulo AA, Leidinger MR, Klesney-Tait JA, Karp PH, Tan P, Wohlford-Lenane C, McCray PB, Meyerholz DK. Heterogeneous expression of the SARS-Coronavirus-2 receptor ACE2 in the human respiratory tract. EBioMedicine. 2020; 60: 102976.

24 Calkovska A, Kolomaznik M, Calkovsky V. Alveolar type II cells and pulmonary surfactant in COVID-19 era. Physiol Res. 2021; 70: S195–S208.

25 Pober JS, Sessa WC. Evolving functions of endothelial cells in inflammation. Nat Rev Immunol. 2007; 7: 803–15.

26 Szmitko PE, Wang CH, Weisel RD, de Almeida JR, Anderson TJ, Verma S. New markers of inflammation and endothelial cell activation: Part I. Circulation. 2003; 108: 1917–23.

27 Varga Z, Flammer AJ, Steiger P, Haberecker M, Andermatt R, Zinkernagel AS, Mehra MR, Schuepbach RA, Ruschitzka F, Moch H. Endothelial cell infection and endotheliitis in COVID-19. Lancet. 2020; 395: 1417–8.

28 Liu F, Han K, Blair R, Kenst K, Qin Z, Upcin B, Wörsdörfer P, Midkiff CC, Mudd J, Belyaeva E, Milligan NS, Rorison TD, Wagner N, Bodem J, Dölken L, Aktas BH, Vander Heide RS, Yin XM, Kolls JK, Roy CJ, Rappaport J, Ergün S, Qin X. SARS-CoV-2 Infects Endothelial Cells. Front Cell Infect Microbiol. 2021; 11: 701278.

29 Øynebråten I, Barois N, Bergeland T, Küchler AM, Bakke O, Haraldsen G. Oligomerized, filamentous surface presentation of RANTES/CCL5 on vascular endothelial cells. Sci Rep. 2015; 5: 9261.

30 Sonmez O, Sonmez M. Role of platelets in immune system and inflammation. Porto Biomed J. 2017; 2: 311-4.

31 Takahashi H, Shiratori M, Kanai A, Chiba H, Kuroki Y, Abe S. Monitoring markers of disease activity for interstitial lung diseases with serum surfactant proteins A and D. Respirology. 2006; 11 **Suppl:** S51-4.

32 Fukaya S, Oshima H, Kato K, Komatsu Y, Matsumura H, Ishii K, Miyama H, Nagai T, Tanaka I, Mizutani A, Katayama M, Yoshida S, Torikai K. KL-6 as a novel marker for activities of interstitial pneumonia in connective tissue diseases. Rheumatol Int. 2000; 19: 223–5.

33 Oyama T, Kohno N, Yokoyama A, Hirasawa Y, Hiwada K, Oyama H, Okuda Y, Takasugi K. Detection of interstitial pneumonitis in patients with rheumatoid arthritis by measuring circulating levels of KL-6, a human MUC1 mucin. Lung. 1997; 175: 379–85.

34 Mitroulis I, Alexaki VI, Kourtzelis I, Ziogas A, Hajishengallis G, Chavakis T. Leukocyte integrins: role in leukocyte recruitment and as therapeutic targets in inflammatory disease. Pharmacol Ther. 2015; 147: 123–35.

35 Hanahan D, Coussens LM. Accessories to the crime: functions of cells recruited to the tumor microenvironment. Cancer Cell. 2012; 21: 309–22.

36 Nozawa H, Chiu C, Hanahan D. Infiltrating neutrophils mediate the initial angiogenic switch in a mouse model of multistage carcinogenesis. Proc Natl Acad Sci U S A. 2006; 103: 12493–8.

37 Pahler JC, Tazzyman S, Erez N, Chen YY, Murdoch C, Nozawa H, Lewis CE, Hanahan D. Plasticity in tumor-promoting inflammation: impairment of macrophage recruitment evokes a compensatory neutrophil response. Neoplasia. 2008; 10: 329–40.

